# Co-circulation of Clade Ia and Ib monkeypox virus in Kinshasa Province, Democratic Republic of the Congo, July - August 2024

**DOI:** 10.1101/2024.09.03.24312937

**Authors:** Tony Wawina-Bokalanga, Prince Akil-Bandali, Eddy Kinganda-Lusamaki, Emmanuel Lokilo, Daan Jansen, Adrienne Amuri-Aziza, Jean-Claude Makangara-Cigolo, Elisabeth Pukuta-Simbu, Rilia Ola-Mpumbe, Cris Kacita, Princesse Paku-Tshambu, Pedro-Henrique L.F. Dantas, Gradi Luakanda, Antoine Nkuba-Ndaye, Meris Matondo, Junior Bulabula, Emmanuel Hasivirwe Vakaniaki, Áine O’Toole, Tessa De Block, Christian Ngandu, Nicole A. Hoff, Nicola Low, Lorenzo Subissi, Sydney Merritt, Jean-Jacques Muyembe-Tamfum, Laurens Liesenborghs, Martine Peeters, Eric Delaporte, Jason Kindrachuk, Anne W. Rimoin, Steve Ahuka-Mundeke, Andrew Rambaut, Dieudonné Mwamba, Koen Vercauteren, Placide Mbala-Kingebeni

**Author notes:** ***Corresponding authors:*** Placide Mbala-Kingebeni, MD, PhD;, Tony Wawina-Bokalanga, MD, PhD (;). Jointly supervised (Koen Vercauteren, Placide Mbala-Kingebeni).

## Abstract

Mpox cases have been reported in nearly all provinces of the Democratic Republic of the Congo as of August 2024. Monkeypox virus positive samples from Kinshasa, collected between July and mid-August 2024, were sequenced using a probe-based enrichment or optimized tiling sequencing protocol. With multiple introductions of both Clade Ia (7/12) and Ib (5/12), marking Kinshasa, and its Limete health zone specifically, as an area with co-circulation of both Clade I, a unique observation illustrating the growing complexity of Clade I mpox outbreaks in DRC.

## Introduction

Mpox, a zoonotic disease caused by monkeypox virus (MPXV), is endemic to Western and Central African countries, with the highest prevalence being in the Democratic Republic of the Congo (DRC) (1). Since the first human case of mpox was identified in the DRC in 1970 (2), efforts to curb the spread of MPXV have been ongoing. However, the number of reported cases has continued to increase over time (3). According to the DRC Ministry of Public Health, as of August 18, 2024, there have been 17,801 suspected cases of mpox reported across nearly all 26 provinces, with 610 deaths, resulting in a case fatality ratio (CFR) of 3.43%.

Before the 2022 multi-country mpox outbreak, MPXV was classified into two major clades: Clade I which is associated with more severe disease and a higher CFR, and Clade II, which is typically associated with milder symptoms (4). Each of these clades is further subdivided: Clade II into IIa and IIb, with IIb variants associated with the 2022 global mpox outbreak driven by human-to-human transmission (5) and Clade I into Ia and Ib, with Ib variants identified in 2023 (6, 7).

While Clade Ia variants remain the most commonly reported in DRC, Clade Ib has been receiving increased attention since its recent emergence in the South Kivu province, eastern DRC. This variant has been associated with sustained human-to-human transmission driven by close contact with infected individuals, including through sexual contact (6).

Recent reports of Clade Ib spreading from its initial outbreak hotspot area, including the North Kivu province (https://virological.org/t/mpox-clade-ib-cases-in-goma/962) as well as internationally (8, 9), have raised international concern. Moreover, the geographic range of Clade I, and predominantly of Clade Ib, is expanding with cases identified in the North Kivu province, DRC (https://virological.org/t/mpox-clade-ib-cases-in-goma/962) as well as in historically non-endemic countries including Burundi, Kenya, Rwanda and Uganda, and has even been imported into countries outside Africa such as Sweden and Thailand (8, 9). Therefore, Africa CDC declared its first public health emergency of continental security (10) and the Word Health Organization declared mpox a public health emergency of international concern for the second time on August 14, 2024 (11). In this context, Kinshasa, the capital city of the DRC, is of particular concern due to high population density of its >17 million people, proximity to Brazzaville, the capital of the Republic of Congo, and international connections via air travel.

Kinshasa is divided into 35 health zones providing healthcare services across its municipalities. Here, we report on mpox cases detected in six health zones of Kinshasa, revealing the co-circulation of both Clade Ia and Ib MPXV variants in the province.

### The Study

As part of routine country-wide mpox surveillance, the Pathogen Genomics laboratory of the Institut National de Recherche Biomédicale (INRB), Kinshasa received vesicle (n=7) and crust (n=4) swab samples, along with one swab of conjunctival secretions from eleven suspected mpox cases in Kinshasa, reported between July and mid-August, 2024. These samples were collected by local surveillance teams, who provided data from each suspected case using the national investigation form. This form includes information on demographic characteristics (age, sex, residence, including health zone and province, profession and nationality), time of onset of clinical symptoms, type of sample and sampling date.

Viral DNA was extracted from 140µL of inactivated swabs resuspended in 1x phosphate buffered saline (PBS) solution using the QIAamp® DNA Mini Kit (Qiagen, Hilden, Germany), following the manufacturer’s instructions. Real-time PCR assays were performed using both Orthopoxvirus- and MPXV-generic primers/probes for mpox diagnosis (12, 13).

We sequenced all samples that tested positive for MPXV. To sequence the full-length MPXV genome, we used either the Comprehensive Viral Research Panel (Twist Biosciences) or the Clade I-optimized tiling sequencing protocol (manuscript in preparation). Sequencing libraries were loaded onto either the MiSeq or GridION sequencer. FASTQ files from the MiSeq were processed using GeVarLi (https://forge.ird.fr/transvihmi/nfernandez/GeVarLi), CZ ID (https://czid.org/), and iVar pipelines. FASTQ files from the GridION were processed using Metatropics (https://github.com/DaanJansen94/nextflow-metatropics-INRB), a Nextflow-driven pipeline for viral pathogen detection and reference-based (GenBank ID:NC_003310) consensus genome generation.

We inferred a Maximum-likelihood phylogenetic tree using IQ-TREE multicore v2.1.4 (14) with the K3Pu+F+I’ model as the best fit, as determined by ModelFinder. Branch support was estimated by ultrabootstrap with 10,000 replicates (15).

All eleven suspected mpox cases were PCR confirmed (100% test positivity ratio) with cycle threshold (Ct) values ranging from 16.81 to 30.59, 45.4% of which were from Limete health zone (5/11) and 18.18% from Kasa-Vubu health zone (2/11) (Figure 1). Each of the remaining health zone: Gombe, Kokolo, Biyela, and Ngiri-Ngiri had 1 confirmed case. Seven of the cases were male and four were female (Table 1).

**Table 1.** Demographic, origin, and clade assignment of mpox cases identified in Kinshasa, DRC. Column percentages are presented in parentheses.

| Age group (years) | Male (n %) | Female (n %) | Health zone | Clade |
| --- | --- | --- | --- | --- |
| 0-10 | 1 (9.09%) | 1 (9.09%) | Kasa-Vubu | Ia |
|  |  |  | Limete | Ib |
| 11-21 | 1 (9.09%) | 2 (18.18%) | Kasa-Vubu | Ia |
|  |  |  | Limete | Ia |
|  |  |  | Limete | Ib |
| 22-32 | 2 (18.18%) | 1 (9.09%) | Limete | Ib |
|  |  |  | Ngiri-Ngiri | Ib |
|  |  |  | Kokolo | Ia |
| 33-43 | 3 (27.28%) | 0 | Limete | Ia |
|  |  |  | Gombe | Ia |
|  |  |  | Biyela | Ia |
| Total | 7 (63.64%) | 4 (36.36%) |  |  |

**Figure 1.**
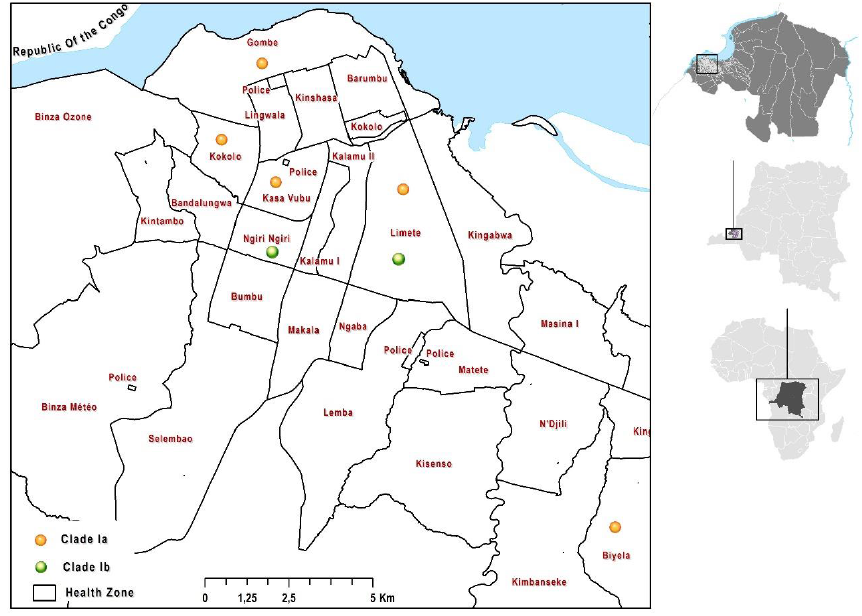
Geographic distribution of mpox confirmed cases, Kinshasa, DRC, July - August 2024. Both Clade Ia and Ib are co-circulating in the health zone of Limete. Mono-circulation pattern of Clade Ia is observed in the following health zones: Gombe, Kokolo, Kasa-Vubu, Biyela and Ngiri-Ngiri. Blue color indicates the Congo River.

MPXV consensus genomes were generated from all eleven cases with horizontal genome coverages ranging from 68.5% to 98.82%, being classified 58.33% (7/12) as Clade Ia and 41.67 (5/12) as Clade Ib MPXV (Table 2). MPXV Clade Ib was detected in age groups 0-10, 11-21, and 22-32 years old (Table 1).

**Table 2.**
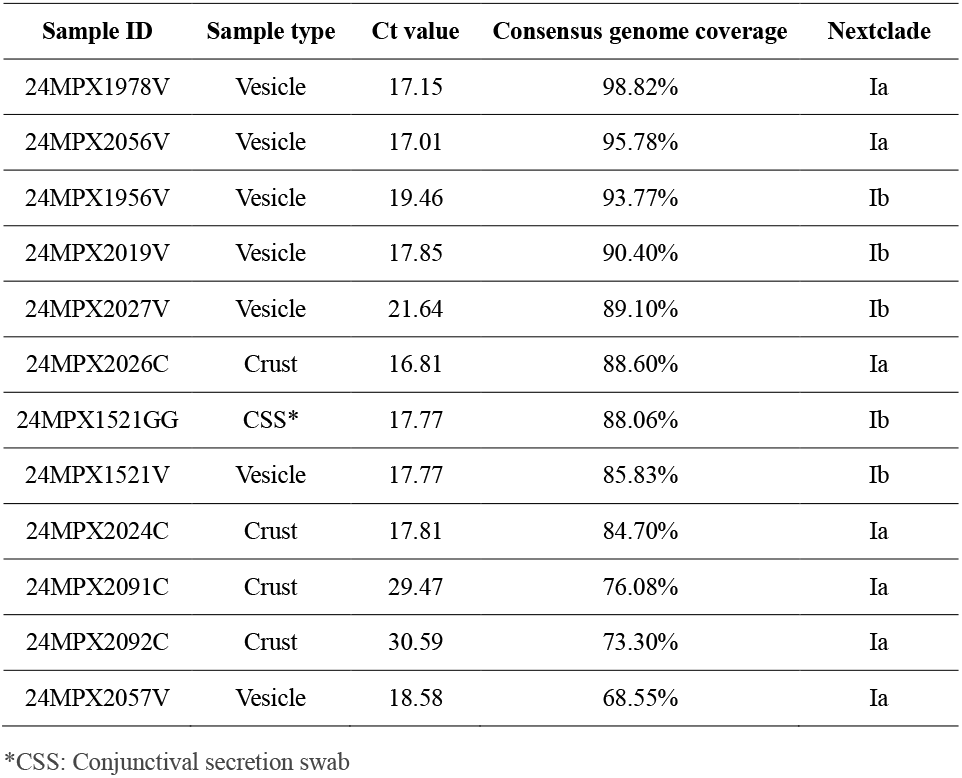
Ct value and genome coverage of MPXV genomes from this study.

Although contact tracing information of mpox confirmed cases was incomplete, the constructed phylogenetic tree (Figure 2) suggests multiple independent introductions of Clade Ia MPXV, and a cluster of Clade Ib, in Kinshasa. While Kinganda-Lusamaki et al. described different groups of Clade Ia lineages co-circulating in five health zones of Kinshasa, including Gombe, Limete, Lemba, Matete, and Nsele (16), we now demonstrate for the first time a co-circulation of both Clade Ia and Ib. This is observed in Kinshasa, and its Limete health zone specifically.

**Figure 2.**
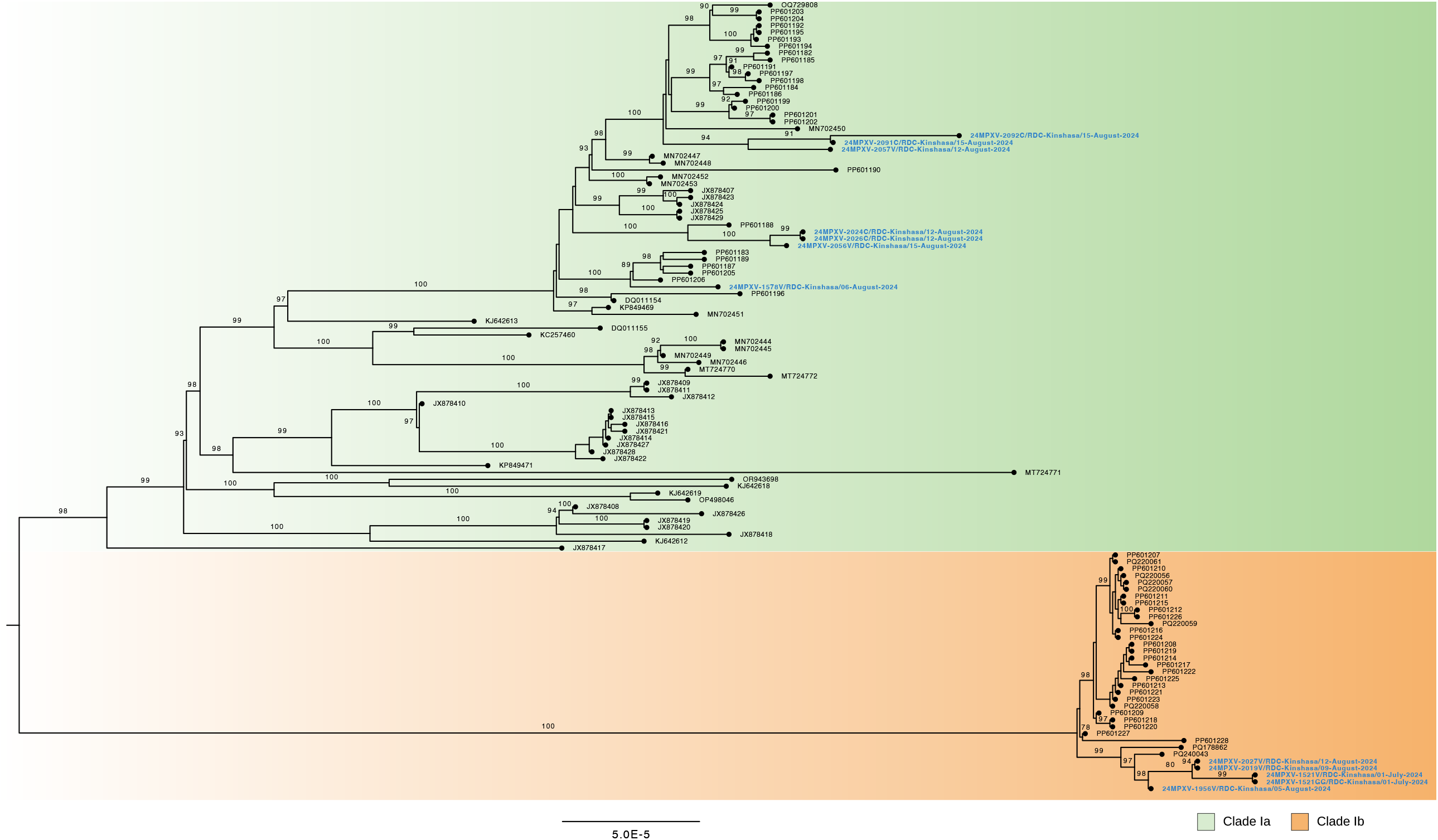
Phylogenetic tree of MPXV sequences (highlighted in blue) from confirmed cases, Kinshasa, DRC, July - August 2024. Sequences are divided into two clades: Clade Ia (green) and Clade Ib (orange).

In this study, mpox cases were confirmed in six of the 35 health zones of Kinshasa. While mpox cases have yet to be reported in the remaining health zones, we consider the possibility of undetected mpox cases in these jurisdictions.

## Conclusions

This study identified co-circulation of two distinct MPXV clades between July and August 2024, Kinshasa, illustrating the complexity of mpox outbreaks in DRC. Ongoing genomic investigations are expected to yield more insights into the circulation of Clade Ia and Ib across different provinces of the DRC. We, therefore, advocate for enhanced surveillance and further epidemiological investigations within the community to better understand and address the factors contributing to mpox outbreaks.

## Data Availability

All data produced in the present study are available upon reasonable request to the authors.

